# Fast intraoperative detection of primary CNS lymphoma and differentiation from common CNS tumors using stimulated Raman histology and deep learning

**DOI:** 10.1101/2024.08.25.24312509

**Authors:** David Reinecke, Nader Maroouf, Andrew Smith, Daniel Alber, John Markert, Nicolas K. Goff, Todd C. Hollon, Asadur Chowdury, Cheng Jiang, Xinhai Hou, Anna-Katharina Meissner, Gina Fürtjes, Maximilian I. Ruge, Daniel Ruess, Thomas Stehle, Abdulkader Al-Shughri, Lisa I. Körner, Georg Widhalm, Thomas Roetzer-Pejrimovsky, John G. Golfinos, Matija Snuderl, Volker Neuschmelting, Daniel A. Orringer

**Author notes:** Corresponding author: David Reinecke, MD Department of Neurosurgery NYU Langone Health, New York University, 550 First Avenue, New York, NY 10016.

## Abstract

**Background:** Accurate intraoperative diagnosis is crucial for differentiating between primary CNS lymphoma (PCNSL) and other CNS entities, guiding surgical decision-making, but represents significant challenges due to overlapping histomorphological features, time constraints, and differing treatment strategies. We combined stimulated Raman histology (SRH) with deep learning to address this challenge. We imaged unprocessed, label-free tissue samples intraoperatively using a portable Raman scattering microscope, generating virtual H&E-like images within less than three minutes. We developed a deep learning pipeline called RapidLymphoma based on a self-supervised learning strategy to (1) detect PCNSL, (2) differentiate from other CNS entities, and (3) test the diagnostic performance in a prospective international multicenter cohort and two additional independent test cohorts. We trained on 54,000 SRH patch images sourced from surgical resections and stereotactic-guided biopsies, including various CNS tumor/non-tumor lesions. Training and test data were collected from four tertiary international medical centers. The final histopathological diagnosis served as ground-truth. In the prospective test cohort of PCNSL and non-PCNSL entities (n=160), RapidLymphoma achieved an overall balanced accuracy of 97.81% ±0.91, non-inferior to frozen section analysis in detecting PCNSL (100% vs. 78.94%). The additional test cohorts (n=420, n=59) reached balanced accuracy rates of 95.44% ±0.74 and 95.57% ±2.47 in differentiating IDH-wildtype diffuse gliomas and various brain metastasis from PCNSL. Visual heatmaps revealed RapidLymphoma’s capabilities to detect class-specific histomorphological key features. RapidLymphoma is valid and reliable in detecting PCNSL and differentiating from other CNS entities within three minutes, as well as visual feedback in an intraoperative setting. This leads to fast clinical decision-making and further treatment strategy planning.

**Importance of the study:** This study introduces RapidLymphoma, a novel self-supervised-based deep-learning model for visual representations to leverage the detection of primary CNS Lymphoma (PCNSL) and differentiation from common and rare CNS entities using intraoperative label-free stimulated Raman histology. While PCNSL is rare, time-critical personalized treatment with fast intraoperative decision-making is needed. In an international multicentric clinical trial, RapidLymphoma first demonstrated its ability to detect and differentiate PCNSL from other CNS lesions with an overall diagnostic balanced accuracy of 97.81% ± 0.91 compared to formalin-fixed paraffin-embedded diagnosis and is non-inferior to frozen section analysis. It provides near real-time intraoperative feedback and guidance to the surgeon, delivering a diagnosis in under three minutes. RapidLymphoma extracts key histomorphological features for detecting and differentiating CNS lesions by utilizing the benefits of intraoperative stimulated Raman histology. This guidance is assisted through a visual prediction heatmap feedback, highlighting critical areas for the surgeon and pathologist.

## Introduction

Primary CNS lymphoma (PCNSL) is responsible for 3-4% of all CNS tumors, suggesting increasing incidence.^1–3^ Diagnostic challenges arise from clinical and imaging similarities between PCNSL and other common CNS entities, such as glial and metastatic tumors. Due to the rapid worsening clinical manifestation in suspected PCNSL, patients often present with neurocognitive deficits, focal neurological deficits, and symptoms of elevated intracranial pressure; therefore, early accurate diagnosis through stereotactic-guided or surgical biopsies is crucial for intraoperative decision-making and rapid further treatment planning.^1^ However, limitations remain in conventional intraoperative histopathological analysis, such as frozen section or cytology smear, in correctly detecting PCNSL and the correct differentiation from non-PCNSL entities, such as adult-type diffuse glioma, metastatic cancer, inflammatory lesions, and other CNS entities with accuracies ranging from 30.8% to 89.6%.^4–7^ Moreover, the diagnostic accuracy rates of final permanent histopathological diagnosis range between 82.2% and 100%, depending on the studied population size and preoperative corticosteroid therapy, remaining hard to diagnose even with immunohistochemistry.^8^ To enable near real-time feedback, reduce prolonged turnaround times, and facilitate fast diagnosis, stimulated Raman histology (SRH) has been introduced as a label-free optical imaging technique for intraoperative histopathological analysis of unprocessed tissue samples to accelerate intraoperative decision-making within minutes.^9–12^ Artificial intelligence-based models (AI), in combination with SRH, have shown that intraoperative streamlined classification is feasible and valid in tumor detection, brain tumor subclassification, and even molecular subtyping of adult-type diffuse gliomas with significantly less need for diagnostic tissue for accurate diagnosis.^13–16^ Moreover, there exists an absence of suitable AI-based methods for detecting PCNSL and differentiating it from other diagnoses. Here, we propose and prospectively test and validate an AI-based approach to enable and accelerate the intraoperative detection of PNCSL and differentiation from other CNS entities through a fully automated SRH whole-slide image analysis pipeline of fresh, unprocessed tissue samples combined with an intraoperative color-coded visualization overlay method to reveal AI-based predictions on whole-slide SRH images.

## Methods

### Study design

The purpose of this study was to (1) develop RapidLymphoma, an intraoperative diagnostic computer vision system that uses SRH and a fully automated AI pipeline to assist the detection, interpretation, and differentiation between PCNSL and non-PCNSL entities based on fresh stereotactic-guided biopsy, surgical, and open biopsy specimens, (2) to perform an international multicenter prospective clinical trial to test the overall balanced accuracy 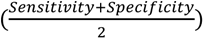, and (3) to compare the overall accuracy of PCNSL cases between frozen section analysis and RapidLymphoma. This study was conducted under institutional review board (IRB) approval from all participating institutions for SRH imaging and data collection. Patient enrollment for intraoperative SRH imaging began in June 2015. Inclusion criteria for intraoperative SRH imaging comprised: (1) suspected central nervous system (CNS) neoplastic/non-neoplastic tissue derived from surgical resection, stereotactic-guided or open biopsy at New York University (NYU), University of Cologne (UKK), Michigan Medicine (UM), and University of Vienna (MUV), (2) subject or caretaker able to give informed consent, and (3) subjects in whom there was additional specimen beyond needed for routine conventional histopathological diagnosis. We trained, validated, and tested an AI-based classification pipeline on patches derived from SRH whole-slide images to provide rapid intraoperative diagnosis of fresh, unprocessed, label-free tissue samples through an intraoperative portable SRH imager. The performance of our developed AI-based pipeline was tested in a prospective international multicenter clinical trial conducted at the prescribed four international tertiary medical centers. A semantic segmentation method was developed and adapted to allow intraoperative visual interpretation of model predictions of SRH whole-slide images for surgeon and pathologist review (Figure 1).^14^

**Figure 1.**
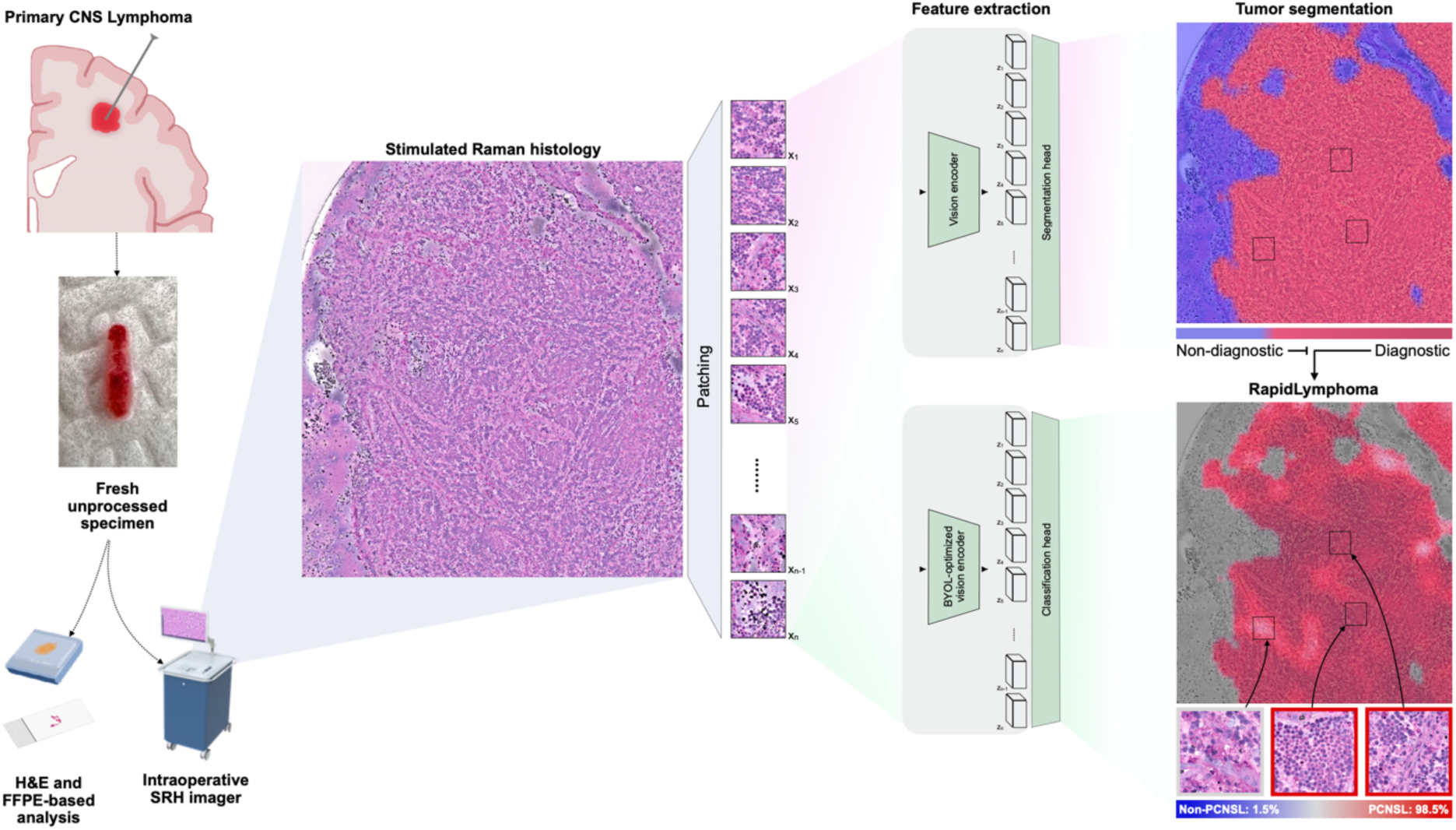
Workflow and Model Implementation for RapidLymphoma. This figure demonstrates the intraoperative workflow and model implementation for detecting primary CNS lymphoma (PCNSL) within minutes using RapidLymphoma. On the left side, the workflow begins with extracting a fresh, unprocessed specimen from a patient with suspected PCNSL during a stereotactic-guided brain biopsy. This specimen is conventionally analyzed using H&E stained, FFPE-based analysis, and an intraoperative Stimulated Raman Histology (SRH) imager. After squeezing on a slide, the SRH imager generates a high-resolution SRH image, highlighting histopathological features. This image is then subjected to patching. Each patch is processed by a segmentation model for diagnostic (tumor, normal brain tissue) and non-diagnostic prediction and is visualized as a heatmap overlay, where non-diagnostic regions are marked in blue and diagnostic tumor regions in red. If tumor regions are detected, our self-supervised vision encoder (BYOL-optimized), called RapidLymphoma, generates latent representations (Z_1_, Z_2_, …, Z_n-1_, Z_n_) of these diagnostic patches. These latent representations are fed into a classification head for our final downstream task to detect PCNSL and differentiate from non-PCNSL brain tumors. The corresponding heatmap overlay excludes non-diagnostic areas (grey) from diagnostic areas (Blue: non-PCNSL, red: PCNSL), with high slide-averaged confidence percentages. Inset images of representative patches illustrate the cellular details corresponding to low-confidence and high-confidence areas for PCNSL, visualizing the model’s predictions.

### Stimulated Raman histology

The whole-slide images used to develop the AI-based pipeline and the prospective international multicenter clinical trial were obtained using a portable fiber-laser-based stimulated Raman scattering (SRS) microscope (NIO Laser Imaging System, Invenio Imaging Inc., Santa Clara, CA, USA). Biomedical tissue is excited by a dual-wavelength fiber laser with a fixed pump beam at 790 nm and a Stokes beam tunable from 1015 nm to 1050 nm. Raman shifts from 2800 cm-1 to 3130 cm-1.^9–11^ Fresh, unprocessed, and label-free tissue samples are prepared on an acrylic slide and squeezed with a cover slip. Images are acquired via beam scanning with spatial sampling of 450nm pixel-1, 1000-pixels per imaged strip, and an imaging speed for 0.4 Mpixel/second/Raman shift sequentially using two specific Raman shifts: 2845 cm-1 and 2930 cm-1. The first shift is responsible for SRS contrast due to CH2 symmetric stretching in fatty acids (lipid-rich areas, CH2 channel). At the same time, the latter shows high contrast in cellular CH3-rich regions (DNA- and protein-rich areas, CH3 channel). A virtual H&E-like image is applied by transforming the raw greyscale SRS images into stimulated Raman histology (SRH) images for visualization purposes on the portable imaging system described previously (Figure 1).^11^

### Image preprocessing

The 16-bit, greyscale CH2 channel images were subtracted pixel-wise from the greyscale CH3 channel images and concatenated to generate a three-channel image (2930 cm-1 minus 2845 cm-1; 2845 cm-1; and 2930 cm-1). As part of the deep learning algorithm development, a 300x300-pixel sliding window technique with a 300-pixel step size was applied to the three-channel image in horizontal and vertical directions to generate non-overlapping image patches matching an eligible input size of the vision encoder in training and inference mode.^13^

### Image training dataset

This study included image datasets with PCNSL and non-PCNSL cases obtained from four SRH imagers at four international neurosurgical institutions: (1) NYU, (2) UKK, (3) UM, and (4) MUV to ensure image data variability, diversity, and generalizability. For training and validation, we used PCNSL and various brain tumor and non-tumor entities as part of the non-PCNSL class to ensure a broad range of visual representations for vision encoder training (Supplemental Figure S1A-C). We employed a pre-trained patch-based whole-slide SRH segmentation model to classify all SRH image patches within each slide as tumor, normal brain tissue, or non-diagnostic (Figure 1).^13,14^ For inclusion in the training dataset, we allowed SRH whole-slide images with up to 90% non-diagnostic patches. This high threshold closely mimics real-world histomorphological image data variability and diversity and fully exploits model capacities during training and inference. It ensured a minimum of 10% diagnostically relevant tissue per slide while maximizing image data utilization. SRH whole-slide images without diagnostic patches or more than 50% normal brain tissue patches were excluded.

### Prospective clinical trial design and image test datasets

We tested the model’s prediction capabilities in a prospective international multicenter clinical trial design with the primary endpoint of overall balanced accuracy 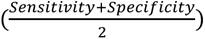. Secondary endpoints included comparing available frozen section or cytology smear analysis of PCNSL cases obtained from the same lesion as SRH with overall accuracy 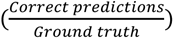 and the model’s ability to learn and distinguish histomorphological features of PCNSL versus non-PCNSL. Prospectively acquired image data, entirely independent from the training cohort, was collected from four neurosurgical institutions: (1) NYU, (2) UKK, (3) UM, and (4) MUV, and ended in June 2024. Clinical trial inclusion criteria were (1) identical to intraoperative SRH imaging, and (2) only diagnostic regions, determined by the pretrained segmentation model, were used(fig). Exclusion criteria comprised: (1) non-diagnostic regions over 90% per SRH image due to substantial blood contamination, cell debris, or artifacts, and (2) broken slide or cover class during tissue preparation.

Based on further fresh-frozen, paraffin-embedded tissue block, and immunohistochemistry analysis, the final histopathological diagnosis was set as appropriate ground truth on the patient-level. The sample size was calculated for an expected accuracy of 96.5%, assuming a null hypothesis of 91.75% accuracy, with 90% statistical power and a two-sided significance level of 5%. The binomial test formula determined a sample size of 160 patient cases.

In addition, the model was tested on two additional independent test cohorts, comprising only (1) IDH-wildtype diffuse gliomas and (2) various brain metastasis to allow performance evaluation on available SRH whole-slide image data beyond the prospective test cohort.

### Deep learning visual representation learning

To enhance the diagnostic accuracy of our intraoperative computer vision classification system for histopathological analysis, we implemented an advanced visual representation learning approach using a convolutional neural network (CNN) as a vision encoder in combination with an adapted self-supervised learning strategy called BYOL - Bootstrap Your Own Latent (Figure 2). This method suits histomorphological learning from stimulated Raman histology (SRH) patch images without negative pairs as needed in other self-supervised contrastive learning algorithms such as SimCLR and SupCon. The algorithm has two main components: an online and a target network. The online network contains a vision encoder *f*_*θ*_, a projection head *g*_*θ*_, and a prediction head *q*_*θ*_, where *θ* are the weights. The target network consists of a vision encoder *f*_*ξ*_ (moving average of *f*_*θ*_), and a projection head *g*_*ξ*_ (moving average of *g*_*θ*_), where *ξ* are the weights of each component. The training process for each iteration is as follows: (1) Sample a minibatch of raw greyscale SRH patch images and apply data augmentations to create two views: *x* and *x*′ of each input patch image. Augmentations include, for example, random cropping, flipping, color jittering, and Gaussian blur. (2) Compute online and target representations:

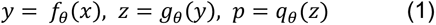

and

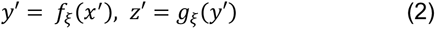

(3) Compute the loss function:

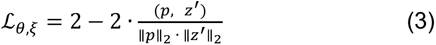

where · denotes the dot product and ||·|| is the L2 normalization. This similarity loss function encourages the online network to predict the target network’s projection, effectively learning invariant representations. (4) Update the online network parameters *θ* only using stochastic optimization to minimize ℒ_*θ*,*ξ*_ as:

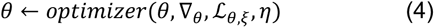

where *optimizer* is an optimizer and *η* is the learning rate. (5) Update the target network parameters *ξ* using an exponential moving average (EMA) as:

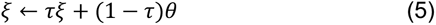

where *τ* is a decay rate. After training, we use the vision encoder *f*_*θ*_ to extract 2048-dimensional feature vectors from SRH patch images.

**Figure 2.**
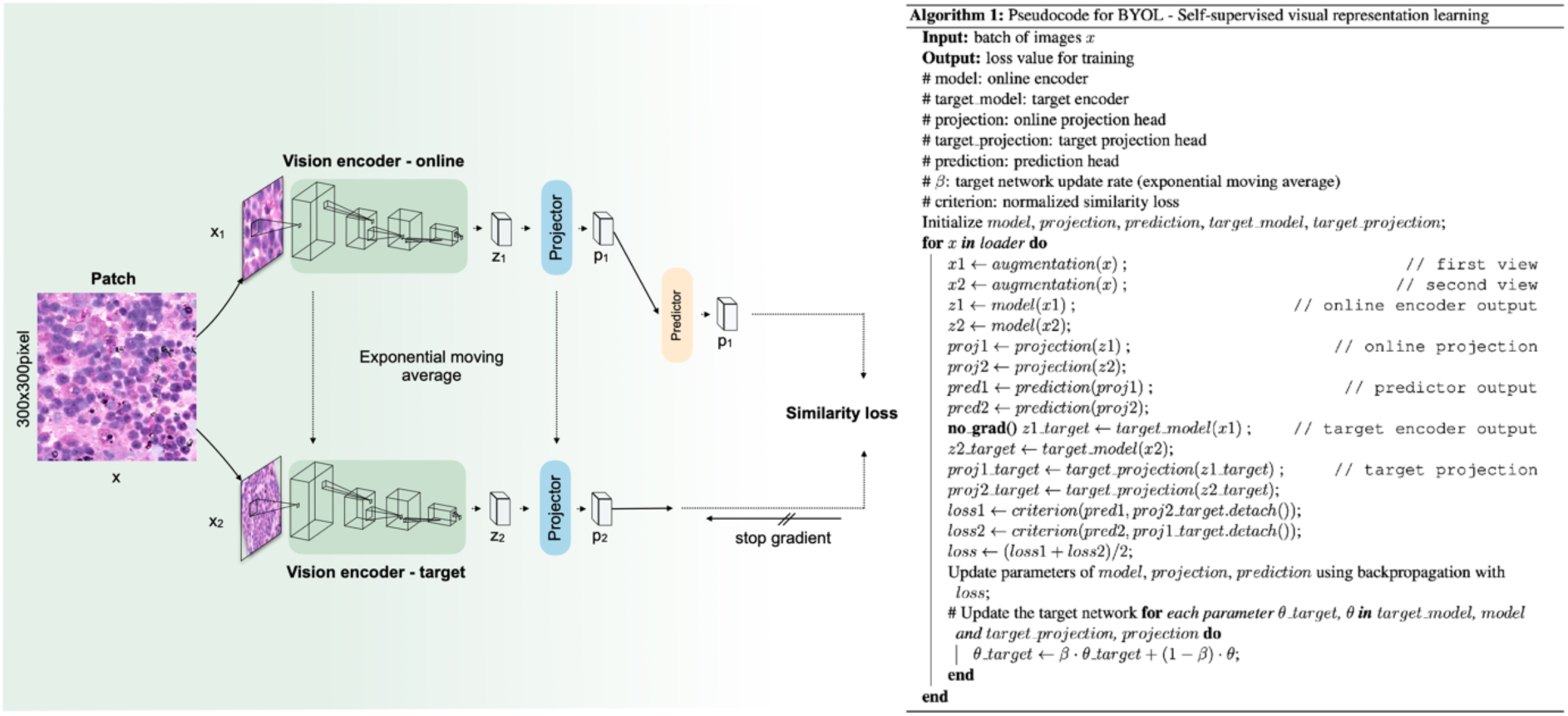
BYOL - Self-Supervised Visual Representation Learning. The figure presents an illustration and pseudocode of the BYOL (Bootstrap Your Own Latent) method for self-supervised visual representation learning. On the left side, the diagram shows the architecture and flow of the BYOL process. It starts with a 300x300 pixel patch of an image X, which is augmented to create two different views, X_1_ and X_2_. These views are fed into two encoders: the online and target encoders. The online encoder processes the first augmented view (X_1_), producing an encoded representation Z_1_, which is then passed through a projector to obtain a projected representation P_1_. A predictor further processes this projected representation to generate the final prediction P_1_. Simultaneously, the target encoder processes the second augmented view (X_2_), producing an encoded representation Z_2_, passed through its projector to obtain a projected representation P_2_. Unlike the online encoder, the gradients from the target encoder are not updated directly; instead, the target encoder is updated using an exponential moving average of the online encoder parameters. The similarity loss is calculated between the online encoder’s predictor output and the target encoder’s projected representation. This loss is used to update the parameters of the online encoder, projector, and predictor via backpropagation. The right side of the figure provides the pseudocode for the BYOL algorithm. The input to the algorithm is a batch of images, and the output is the loss value used for training guidance and hyperparameter tuning.

Our experiment utilized a ResNet50 vision encoder without pretraining and the described self-supervised learning approach. We trained on 300x300 pixel SRH image patches with a strong augmentation strategy, including random horizontal and vertical flips, sharpness adjustments, Gaussian blur, Gaussian noise, auto contrast, solarization, and random cropping. The model was trained for 270 epochs with a batch size of 128 images using the AdamW optimizer (learning rate 0.001) and a cosine learning rate decay with linear warmup. A five-fold hold-out cross-validation strategy was performed every two epochs with balanced sampling across classes to ensure unbiased evaluation of the model’s performance.

### Linear downstream task for binary classification

In this downstream task, we leverage a pre-trained visual encoder for the binary classification of PCNSL and non-PCNSL cases. Following training our visual encoder *f(⋅)* through self-supervised learning, we freeze its parameters and implement a single linear layer for classification. The output of this downstream classification layer can be described as follows:

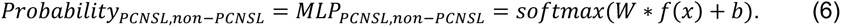

Softmax represents the activation function that generates a probability distribution over the two classes between 0 and 1. *W* denotes the weight matrix, and *b* represents the bias vector. We employ the cross-entropy loss function to train this downstream classification layer:

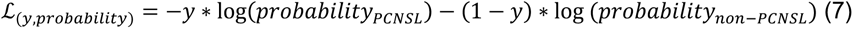

In this formula, y represents the true label (1 for PCNSL, 0 for non-PCNSL), while *probability_PCSNL_* and *probability_non-PCNSL_* are the model’s predicted probabilities for the respective classes. This downstream approach enables simple but efficient binary classification based on the features extracted by the pre-trained visual encoder. We trained the linear layer on the same dataset used for the vision encoder for five epochs without any additional strong augmentation strategy and with a batch size of 128 using the AdamW optimizer (learning rate 0.01) and a cosine learning rate decay with linear warmup.

### Semantic probability segmentation and visual heatmap generation

We implemented a semantic probability segmentation approach to generate heatmap overlays over the SRH image to enhance the interpretability of our developed computer vision system and provide visual feedback to surgeons and pathologists. This technique allows for the localization of regions within the SRH whole-slide images most indicative of PCNSL or non-PCNSL characteristics. For probability heatmap overlay generation, we implemented a function that performs a sliding window approach over the input whole-slide image, applying the pretrained tumor segmentation model and our developed model to each raw greyscale SRH image patch. A defined patch size of 300x300 pixels and a step size of 100 pixels is used for the function to systematically move this window across the entire image input in both *x* x and *y* directions. For each position (*x*, *y*) we extract a 300x300 pixel patch, which is processed first for diagnostic tumor segmentation and then for classification. The output for the segmentation model heatmap is

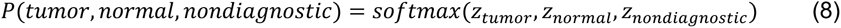

are where *P* is the probability and *z* is the logit output of the segmentation model. The softmax function puts all three classes into one probability range between 0 and 1. For heatmap generation, an empty heatmap array is initialized of the same size as the whole-slide image with an additional dimension for each class. The patches classified as diagnostic tumor are only used now for probability predictions in the final heatmap using the sigmoid function to allow value-depending heatmap coloring with

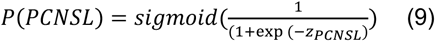

where each patch contains the probability of PCNSL class, but only for areas classified as a tumor. For color-coded heatmap visualization, probability values are extracted from the previously calculated PCNSL class heatmap array, where red indicates high probability, light/white for uncertainty, blue for low probability for the PCNSL class, and due to the binary classification, and vice versa for the non-PCNSL class. Finally, the overlay is created using a weighted sum of the original whole-slide image and the heatmap with a defined transparency factor to allow intuitive interpretation of the class probability predictions. Additionally, the probability predictions are summed over the SRH whole-slide image to give a final whole-slide-based prediction for each class with

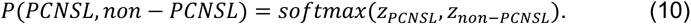

### Ablation studies

We conducted a primary ablation experiment to evaluate the significance of crucial training strategy choices in our approach. This experiment used the same training dataset and the prospective international multicenter test cohort. We compared supervised cross-entropy versus our self-supervised learning strategy for visual representation learning. Both approaches utilized a ResNet-50 model without any pre-trained weights. The cross-entropy model was trained in a conventional supervised manner, while the other model employed our adapted self-supervised learning strategy. All experiments used the same batch size (128), optimizer (AdamW), and learning rate schedule (cosine decay with linear warmup) described in our central methodology.

### Statistics and reproducibility

We employed a comprehensive set of statistical metrics to evaluate our model’s performance against the final histopathological diagnosis as ground truth. We calculated accuracy, balanced accuracy, sensitivity, specificity, area under the receiver operating characteristic curve (AUROC), and F1 score in percentage, along with their standard deviations and 95% confidence intervals. This provided a robust assessment of the model’s reliability and consistency. All whole-slide corresponding patch softmax probabilities were aggregated and summed using the mean for level-dependent (patient and slide) metric evaluation. We utilized two dimensionality reduction techniques to visualize the learned visual patch representations: t-distributed stochastic neighbor embedding (t-SNE) and pairwise controlled manifold approximation (PaCMAP) to offer advantages in maintaining both local and global structures, which are important in revealing different subclasses.

### Computational hardware and software

All images used for training, validation, and testing were processed using a single Nvidia A100 80-gigabyte graphical processing unit (GPU) accelerator with a 4 x 16-gigabyte central processing unit (CPU) with our custom Python-based (version 3.10.13) RapidLymphoma package. We used the Pydicom package (version 2.4.3) to process the SRH images from the NIO Laser Imaging System. All archived post-processed images have been stored as 16-bit TIFF images on a high-performance cluster for further tasks using the tifffile package (version 2023.9.26). All models were trained using the New York University BigPurple hybrid cluster. BigPurple is a high-performance cluster environment that aligns with HIPAA privacy standards. All AI-based models were trained on a single Nvidia A100 80-gigabyte GPU accelerator. All custom code for training and evaluation can be found in our open-source RapidLymphoma repository. Our model was implemented in PyTorch (version 2.1.2). We used an untrained customized ResNet-50 model from torchvision (0.10.10). Scikit-learn (version 1.4.0) was used to compute performance metrics on model predictions at training and inference.

## Results

We developed RapidLymphoma, an intraoperative diagnostic computer vision system designed to differentiate between primary CNS lymphoma (PCNSL) and non-PCNSL entities using stimulated Raman histology (SRH) imaging of fresh surgical tissue samples. This system employs a fully automated AI-based pipeline to assist fast intraoperative interpretation of SRH whole-slide images obtained from stereotactic-guided biopsies, surgical resections, and open biopsies. The development of RapidLymphoma involved several key components, including our tumor segmentation model, our self-supervised learning approach to train a robust vision encoder on SRH patch images, a downstream linear classification task to distinguish between PCNSL and non-PCNSL cases, and a semantic probability segmentation technique to generate interpretable visual heatmaps for surgeons and pathologists (Figure 1). To validate the performance of RapidLymphoma, we conducted an international multicenter prospective clinical trial across four tertiary medical centers: (1) NYU, (2) UKK, (3) UM, and (4) MUV.

### Prospective clinical trial demographics

We tested on a prospective cohort of 160 patients, comprising 88 males and 72 females (Figure 3A-C). The study utilized SRH whole-slide images, ranging from 1 to 17 per patient. Within this cohort, PCNSL accounted for 25 patients (15.62%), while non-PCNSL diagnoses were observed in 135 cases (84.38%). Among the non-PCNSL diagnoses, adult-type diffuse glioma with IDH-wildtype status emerged as the most prevalent, representing 52.50% of all cases. This was followed by metastases (12.50%) and adult-type diffuse glioma with IDH-mutant status (10.62%). Each of these diagnostic categories encompassed various subdiagnoses, reflecting the heterogeneity of these categories. The remaining cases consisted of rare tumors and non-tumorous lesions. Regarding the diagnostic procedures employed, stereotactic-guided biopsies were the primary diagnostic tool for PCNSL cases, accounting for 16.88% of all procedures. In contrast, surgical resection was the predominant approach for non-PCNSL cases, representing 55.62% of the procedures. Open biopsy was utilized in two cases (1.25%). Also, the study included primary and recurrent brain lesions, with primary diagnoses constituting the majority at 93.75% of cases. Demographic analysis of the cohort revealed that most patients were White (53.12%), while Asian and Black or African American each represented less than 10% of the study population.

**Figure 3.**
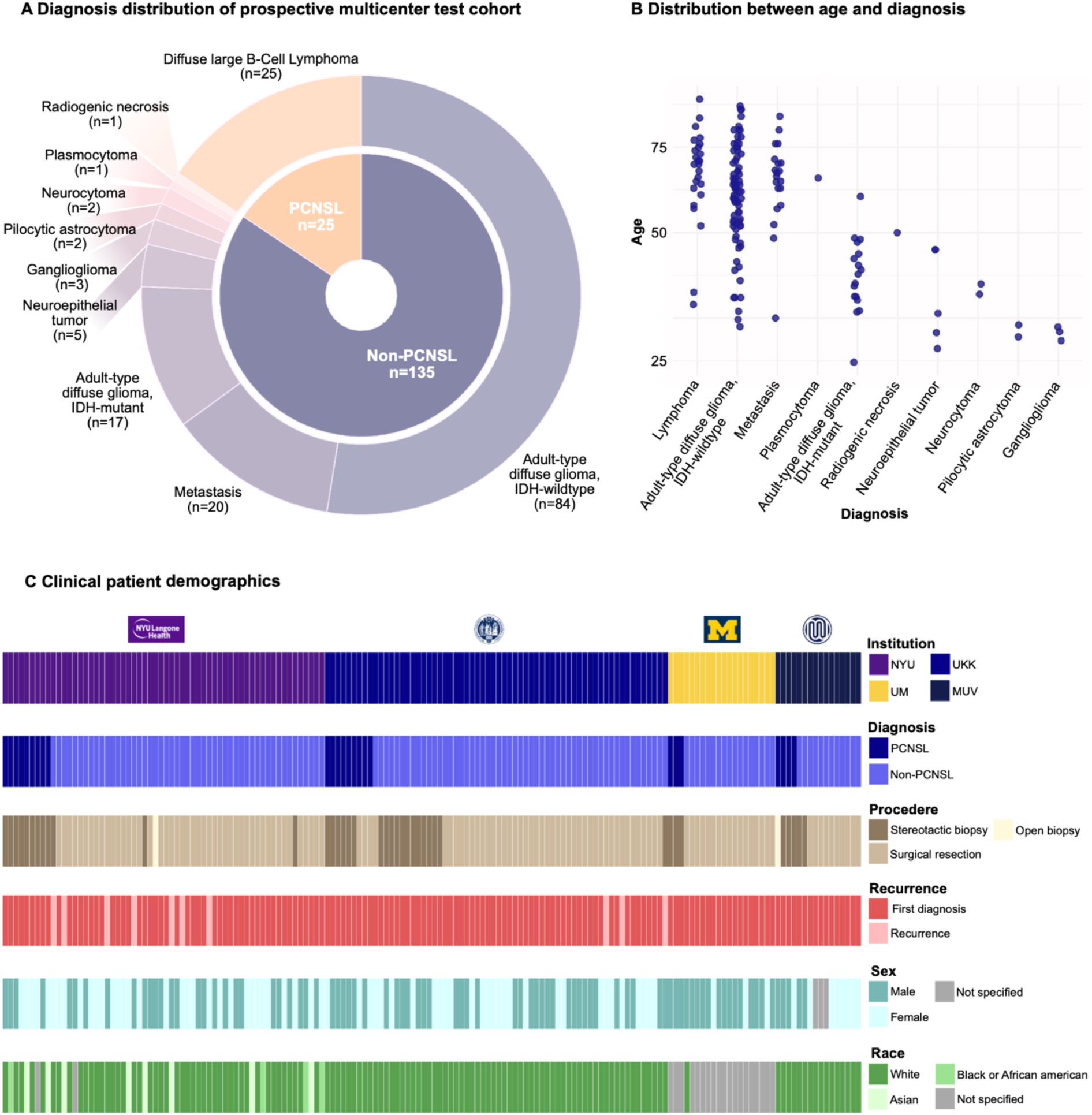
Prospective international multicenter testing cohort demographics. The figure provides an overview of the demographics and clinical characteristics of our prospective international multicenter cohort used to evaluate RapidLymphoma. **A.** categorizes the cohort into PCNSL (Primary Central Nervous System Lymphoma) and non-PCNSL cases. Out of 160 total cases, 25 are identified as PCNSL, while 135 are non-PCNSL, which includes a variety of other diagnoses to ensure that the classifier is tested on various CNS entities. **B.** illustrates the age distribution of patients across different diagnoses to reveal age-related correlations. **C.** Stacked bar plots further break down the cohort based on several demographic and clinical variables. The patients come from four major institutions: New York University (NYU), University Hospital Cologne (UKK), University of Michigan (UM), and Medical University of Vienna (MUV). This geographic and institutional diversity adds robustness to the evaluation by incorporating different clinical practices and patient populations.

### Overall diagnostic performance in the prospective clinical trial

The analysis was conducted at three levels: patient (n=160), slide (n=213), and patch (n=43083). The model demonstrated high overall balanced accuracy across all levels: 97.81% ± 0.91 (95% CI: 95.96-99.29) at the patient-level, 97.21% ± 1.11 (95% CI: 94.71-99.03) at the slide-level, and 90.88% ± 0.16 (95% CI: 90.58-91.20) at the patch-level (Figure 4B). Specificity and sensitivity were consistently high, with 100% ± 0 (95% CI: 100.00-100.00) and 95.68% ± 1.70 (95% CI: 92.14-98.56) for patient-level and 98.66% ± 1.40 (95% CI: 95.52-100.00) and 95.84% ± 1.58 (95% CI: 92.57-98.64) for slide-level. AUROC and F1 scores revealed high performance with 99.71% ± 0.25 (95% CI: 99.13-100.00) and 97.78% ± 0.89 (95% CI: 95.91-99.28) at the patient-level (Figure 4A) and 98.76% ± 1.02 (95% CI: 96.45-99.95) and 97.55% ± 0.92 (95% CI: 95.59-99.02) at the slide-level. The patch-based softmax probability distribution showed a clear separation between PCNSL and non-PCNSL cases (Figure 4E), correctly identifying all 25 PCNSL cases without false negatives at the patient level and accurately classifying 129 out of 135 non-PCNSL cases with six false positive cases. On slide-level, 68 PCNSL slides with six false positives and 138 non-PCNSL slides with one false negative were classified (Figure 4D, Figure 5, Supplemental Figure S2 and S3). We conducted subgroup analyses with age, gender, race, procedure, and institution to evaluate performance across various clinical demographics. The model consistently demonstrated balanced accuracies and AUROC values with over 95% (Supplemental Figure S4 and S5). A closer look at the dimensionality reduction graphs (PaCMAP) objectified a separation between the PCNSL and non-PCNSL class with clustering of patches belonging to the same slide and patient, as well as in the non-PCNSL to the same entity (Figure 6).

**Figure 4.**
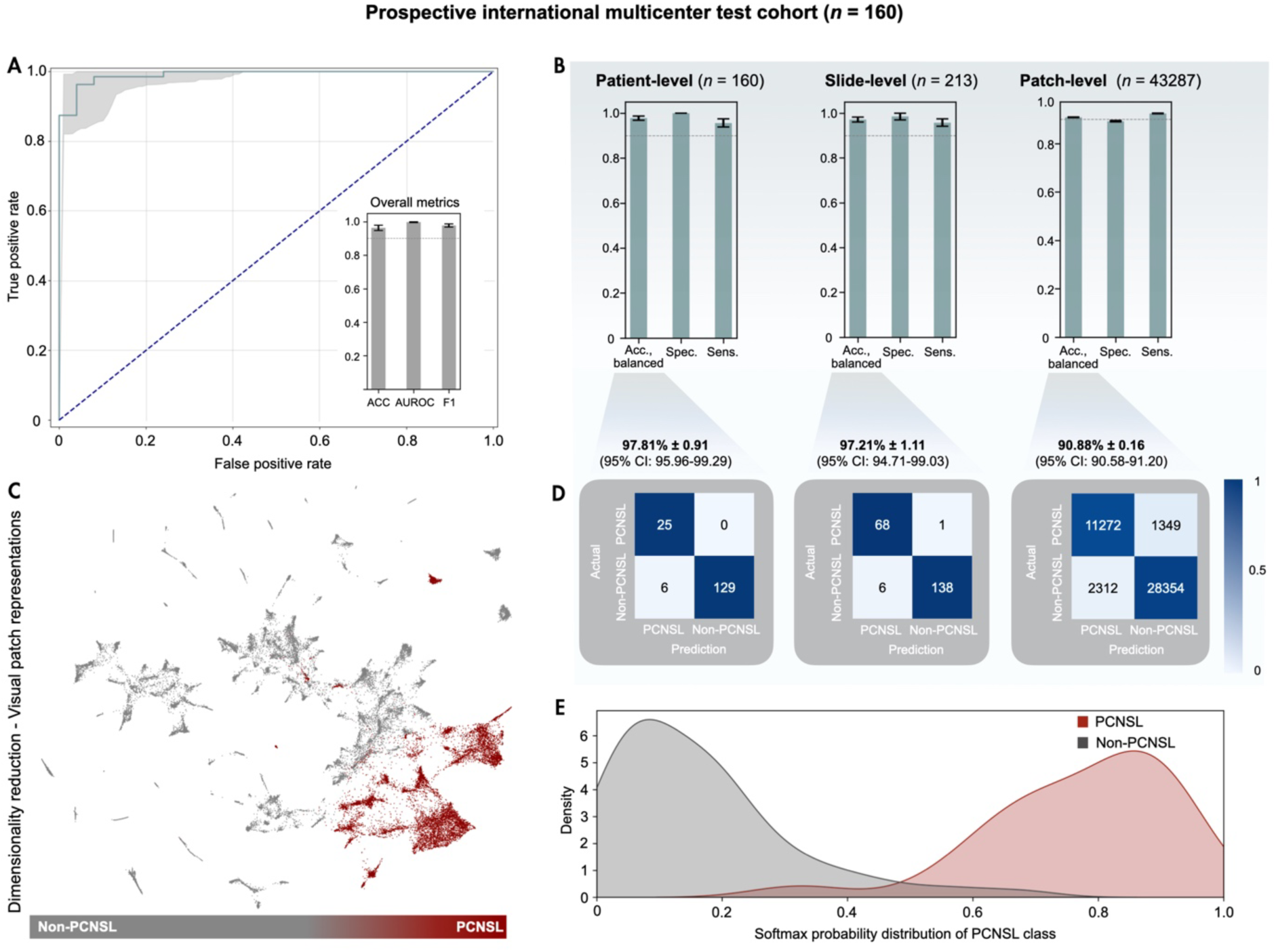
RapidLymphoma Classification Performance. **A.** The ROC curve for the prospective multicenter test cohort (n = 160) illustrates a high true positive rate with a low false positive rate to distinguish between the PCNSL and non-PCNSL class. The inset bar plot displays performance metrics: Accuracy (ACC), area under the receiver operating characteristic curve (AUROC), and F1 score show high diagnostic performance. **B.** This panel compares classification performance across three different levels: patient-level (n = 160), slide-level (n = 213), and patch-level (n = 43287). The patient-level classification shows the highest accuracy and balanced performance, followed by the slide-level and patch-level classifications. These plots highlight the consistency of model performance across different granularities of the data. **C.** A PaCMAP (Pairwise Controlled Manifold Approximation) plot illustrates the clustering of primary CNS lymphoma (PCNSL, red) and non-PCNSL entities (grey) with shared local and global histomorphological key features in a two-dimensional space, providing a visual representation of the strong separation achieved by the classifier. **D.** Confusion matrices at patient, slide, and patch-level detail the number of true positives, true negatives, false positives, and false negatives. Each matrix includes the classification level’s overall accuracy and 95% confidence intervals (CI). **E.** A density plot shows the softmax probability distribution for the PCNSL class. The distribution is presented for both the PCNSL and non-PCNSL class. The plot reveals a clear separation in the predicted probabilities, with PCNSL class probabilities clustering towards higher values, indicating high confidence in the classifier’s predictions for the PCNSL class, while the non-PCNSL class shows a density peak within very low class probabilities.

**Figure 5.**
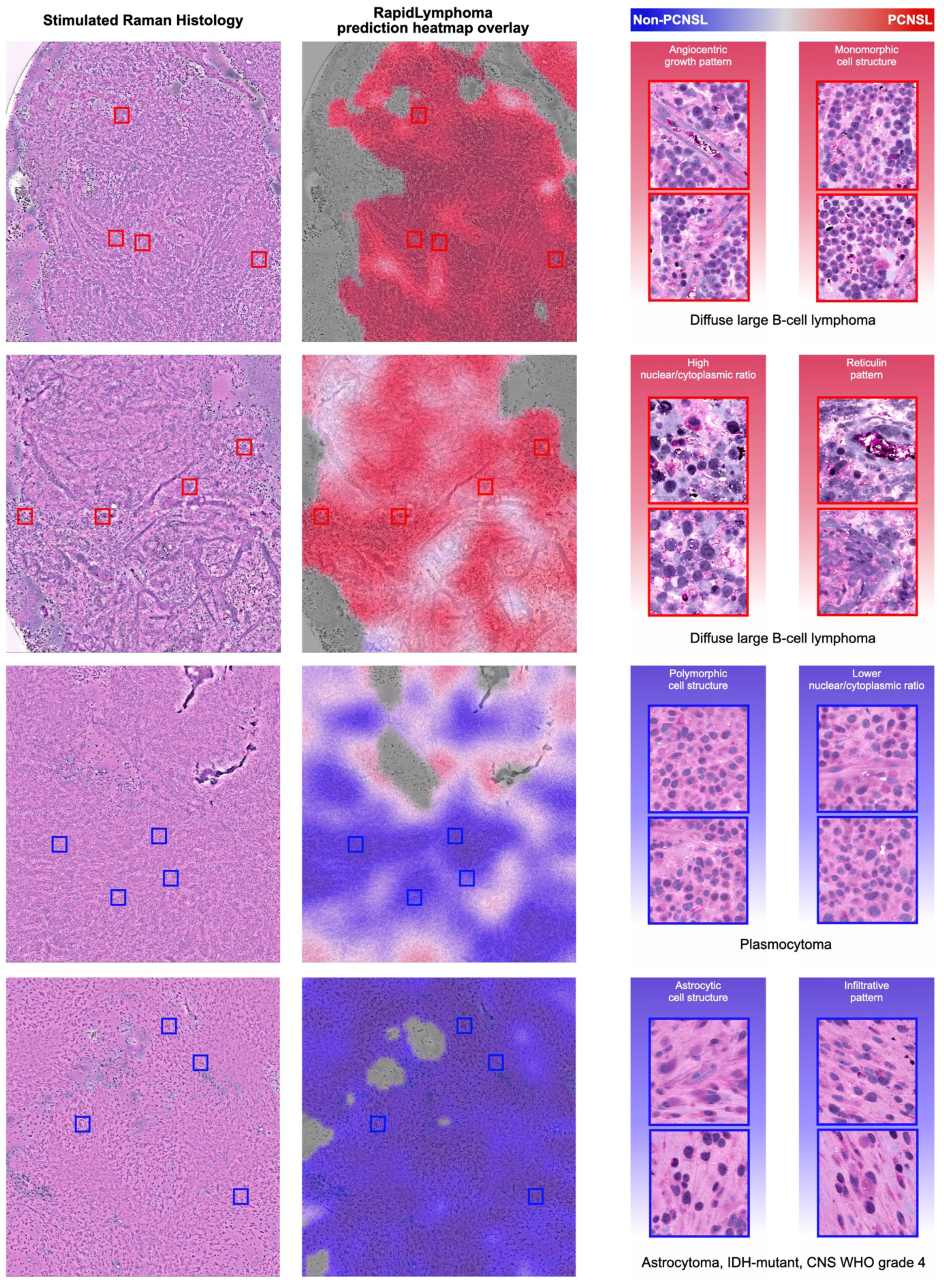
Correctly Predicted Whole Slide Images from the Prospective Testing Cohort. Correctly predicted whole slide images from the prospective testing cohort demonstrate the model’s effectiveness in distinguishing between PCNSL and non-PCNSL tissues. The figure is organized into three main columns: Stimulated Raman Histology (SRH) images, RapidLymphoma’s prediction heatmap overlays, and detailed image patch comparisons. Red indicates high confidence for PCNSL, blue indicates non-PCNSL, and white/grey indicates low confidence. The detailed image patches on the right side from the first two PCNSL slides demonstrate an angiogenic growth pattern, high nuclear/cytoplasmic ratio, monomorphic cell structure, and reticulin pattern. In contrast, patches from the other two non-PCNSL slides illustrate polymorphic cell structure, lower nuclear/cytoplasmic ratio, and astrocytic cell structure. The first two rows are featured by a diffuse large B-cell lymphoma, the other two by a plasmocytoma, and an astrocytoma, IDH-mutant CNS WHO grade 4. Image quality impacts RapidLymphoma’s predictions with lower confidence, resulting in white/grey patch regions.

**Figure 6.**
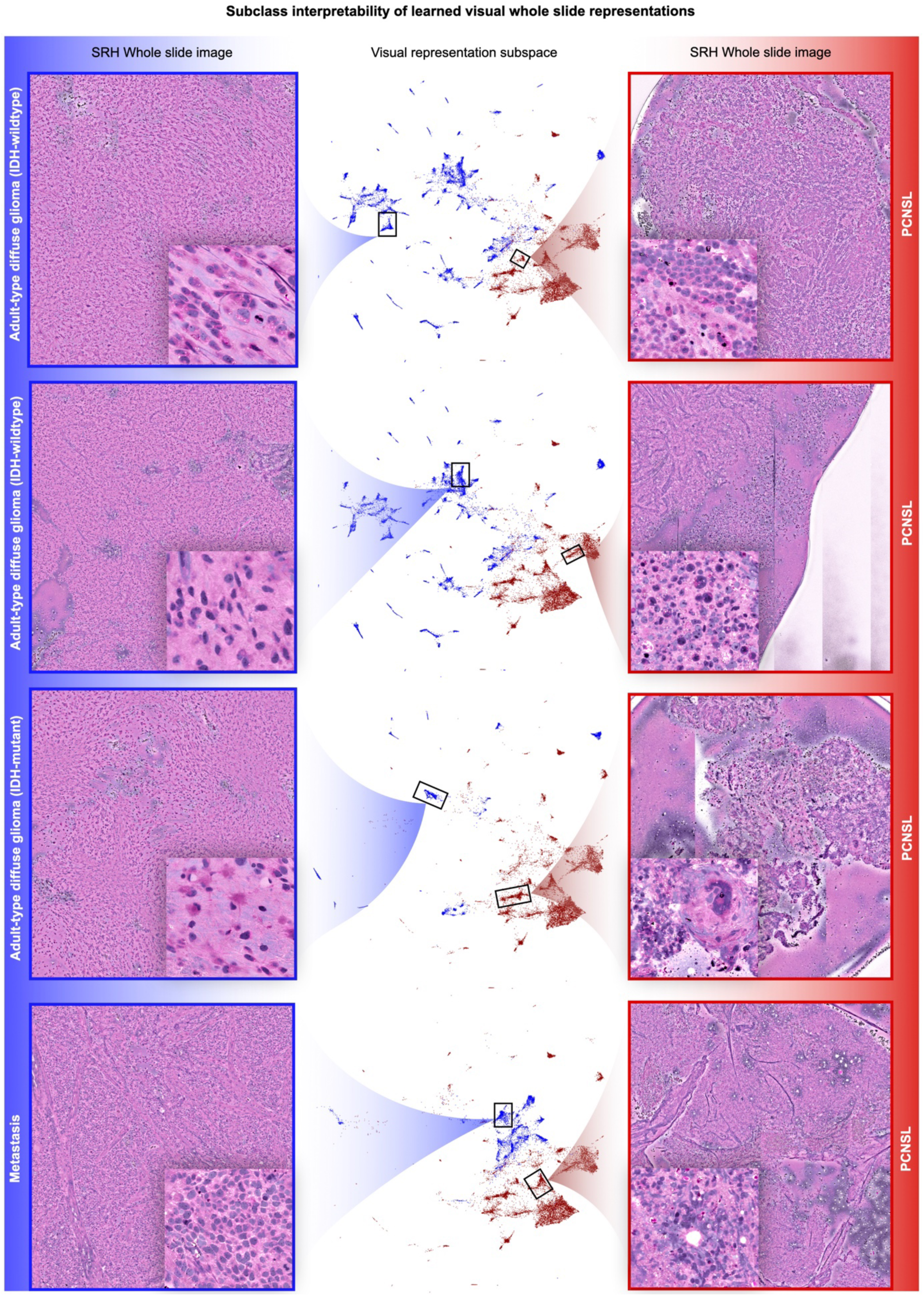
Subclass Interpretability of Learned Visual Whole Slide Representations. Here, we demonstrate the subclass interpretability of the RapidLymphoma model’s learned visual whole slide representations, focusing on different histopathological subtypes. The figure is organized into three columns: SRH whole slide images, visual representation subspaces, visualized using PaCMAP (Pairwise Controlled Manifold Approximation Projection), and SRH whole slide images for PCNSL. The left column shows representative SRH whole-slide images for adult-type diffuse glioma and metastasis. Each image includes a detailed inset highlighting specific histopathological features. These slides are framed in blue to indicate the non-PCNSL category visible in PaCMAP. Each point represents a patch from the SRH images, color-coded to differentiate between PCNSL (red) and non-PCNSL (blue) regions. The plot shows how the model organizes these patches into distinct whole slide clusters, with blue points corresponding to non-PCNSL subtypes and red points to PCNSL. The clear separation between the clusters indicates the model’s capacity to distinguish between different histopathological features and cluster them according to similar patches from the same whole slide image in the representation space. The right column features SRH whole slide images for PCNSL (red), which are also accompanied by detailed insets that emphasize the characteristic features of PCNSL. This figure demonstrates how it leverages these differences to organize the visual representations in a structured manner on slide-level. The interpretability provided by this visualization allows for a deeper understanding of the model’s decision-making process.

### Performance comparison to frozen section/cytology smear in the prospective clinical trial

The performance of RapidLymphoma detection for PCNSL cases was compared to conventional frozen section analysis. No cytology smear analysis was performed. While tissue for SRH was available for all 25 PCNSL cases, frozen section analysis was only performed in 18 cases. Among these, four cases were incorrectly diagnosed by frozen section. Specifically, two cases were misdiagnosed as metastasis, one case as a glial tumor, and one case was misidentified as an epithelioid tumor, leading to an accuracy of 77.77% (14/18).

### Diagnostic performance on two additional external test cohorts

In addition to the prospective cohort, we evaluated two independent test cohorts: PCNSL versus IDH-wildtype diffuse glioma and brain metastasis. The first cohort comprised 420 patients, including 25 patients of PCNSL. The mean age was 62.93 years with a slight male predominance (59.28% male, 40.72% female). Racial distribution, available for 189 patients, indicated most White (88.89%), followed by Asian (5.82%) and Black or African American (5.29%) patients. Most non-PCNSL cases were glioblastoma, IDH-wildtype, CNS WHO grade 4 (94.94%). Rare diagnoses (5.06%) comprised diffuse midline glioma, gliosarcoma, diffuse hemispheric glioma, and anaplastic glioma NOS. Surgical resection was the primary procedure, while all cases were newly diagnosed.

RapidLymphoma demonstrated a balanced accuracy of 95.44% ± 0.74 (95% CI: 93.97-96.84) with an AUROC of 98.87% ± 0.46 (95% CI: 97.87-99.67) at the patient-level. At the slide-level (n=1772), the model maintained a balanced accuracy of 94.21% ± 0.82 (95% CI: 92.37-95.47) with an AUROC of 97.43% ± 1.12 (95% CI: 94.86-98.94) (Figure 7A,C,E). The second test cohort focused on distinguishing PCNSL from brain metastases, comprising 59 patients with a mean age of 65.46 years, also showing a slight male predominance (54.24% male, 45.76% female). Of the 55 patients with available racial data, 89.09% were White, 5.45% were Black or African American, and 5.45% were Asian. The cohort included 34 cases of metastasis, with adenocarcinoma of the lung being the most common metastatic tumor type (79.41%). Further diagnoses (20.59%) comprised urothelial carcinoma, squamous cell carcinoma, melanoma, adenoid cystic carcinoma, and adenocarcinoma of the rectum. Surgical resection was the primary procedure, and all cases were newly diagnosed. At the patient-level, RapidLymphoma achieved a balanced accuracy of 95.57% ± 2.47 (95% CI: 90.00-100.00) and an AUROC of 99.88% ± 0.19 (95% CI: 99.30-100.00). The slide-level analysis (n=152) revealed a balanced accuracy of 95.74% ± 1.60 (95% CI: 92.51-98.68) and an AUROC of 97.49% ± 1.39 (95% CI: 94.33-99.56) (Figure 7B,D,F).

**Figure 7.**
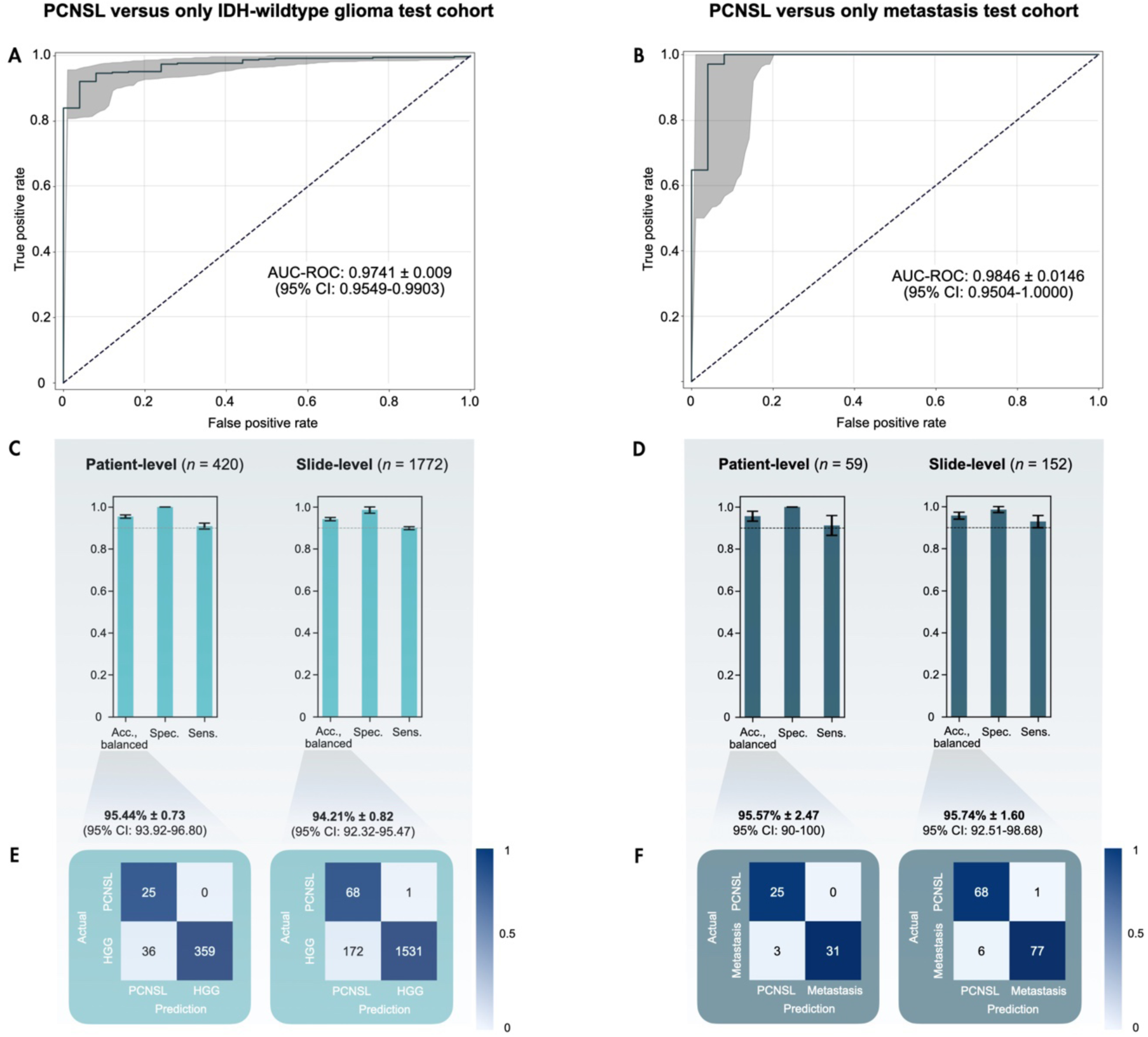
RapidLymphoma performance evaluation of high-grade glioma and metastasis. The figure illustrates the performance of RapidLymphoma in distinguishing primary central nervous system lymphoma (PCNSL) from two common differential diagnoses: adult-type diffuse glioma (IDH-wildtype) and metastasis. The data was derived from independent and external test cohorts to ensure further robust evaluation. On the left side, performance metrics are presented for HGG, and on the right, for metastasis. Due to the rarity and limited availability of lymphoma cases, the same stimulated Raman histology (SRH) images from the prospective cohort were utilized to generate the metrics. These presented test cohorts are independent and external from the primary prospective test cohort. This approach was adopted to provide a more accurate depiction of the classifier’s performance against the typical differential diagnoses such as IDH-wildtype glioma (Glioblastoma, diffuse midline glioma, gliosarcoma, and diffuse hemispheric glioma) and metastasis (Melanoma, various adenocarcinoma, squamous cell carcinoma, mamma carcinoma, and neuroendocrine carcinoma). **A.** A Receiver Operating Characteristic (ROC) curve represents the classifier’s ability to differentiate between PCNSL and adult-type diffuse glioma (IDH-wildtype), indicating high diagnostic performance. **B.** This plot demonstrates the classifier’s performance in distinguishing PCNSL from brain metastasis with another ROC curve, suggesting excellent classification capability. **C.** This panel shows the performance metrics for PCNSL versus adult-type diffuse glioma (IDH-wildtype). The bar charts present the balanced accuracy, specificity, and sensitivity at the patient level (n = 437) and slide level (n = 1736). The classifier exhibits high performance across all metrics, indicating its reliability in distinguishing PCNSL from high-grade glioma. **D.** The performance metrics for PCNSL versus metastasis are depicted in this panel. Like panel C, these bar charts present balanced accuracy, specificity, and sensitivity at the patient level (n = 59) and slide level (n = 149). The results consistently show high performance, underscoring the classifier’s effectiveness in differentiating PCNSL from brain metastasis. **E.** The confusion matrices for PCNSL versus adult-type diffuse glioma (IDH-wildtype; HGG) are shown in this panel to reveal false positive and negative predictions. **F.** The confusion matrices for PCNSL versus metastasis are illustrated in this panel.

### Ablation studies

To evaluate our algorithm against other common learning strategies, we compared RapidLymphoma to a cross-entropy (CE) experiment conducted on the prospective international multicenter test cohort. The ablation studies revealed distinct patterns in the visual patch representation space for the RapidLymphoma self-supervised method and the CE supervised experiment. Both strategies were able to cluster the PCNSL and non-PCNSL class (Figure 8A and Supplemental Figure S6).

**Figure 8.**
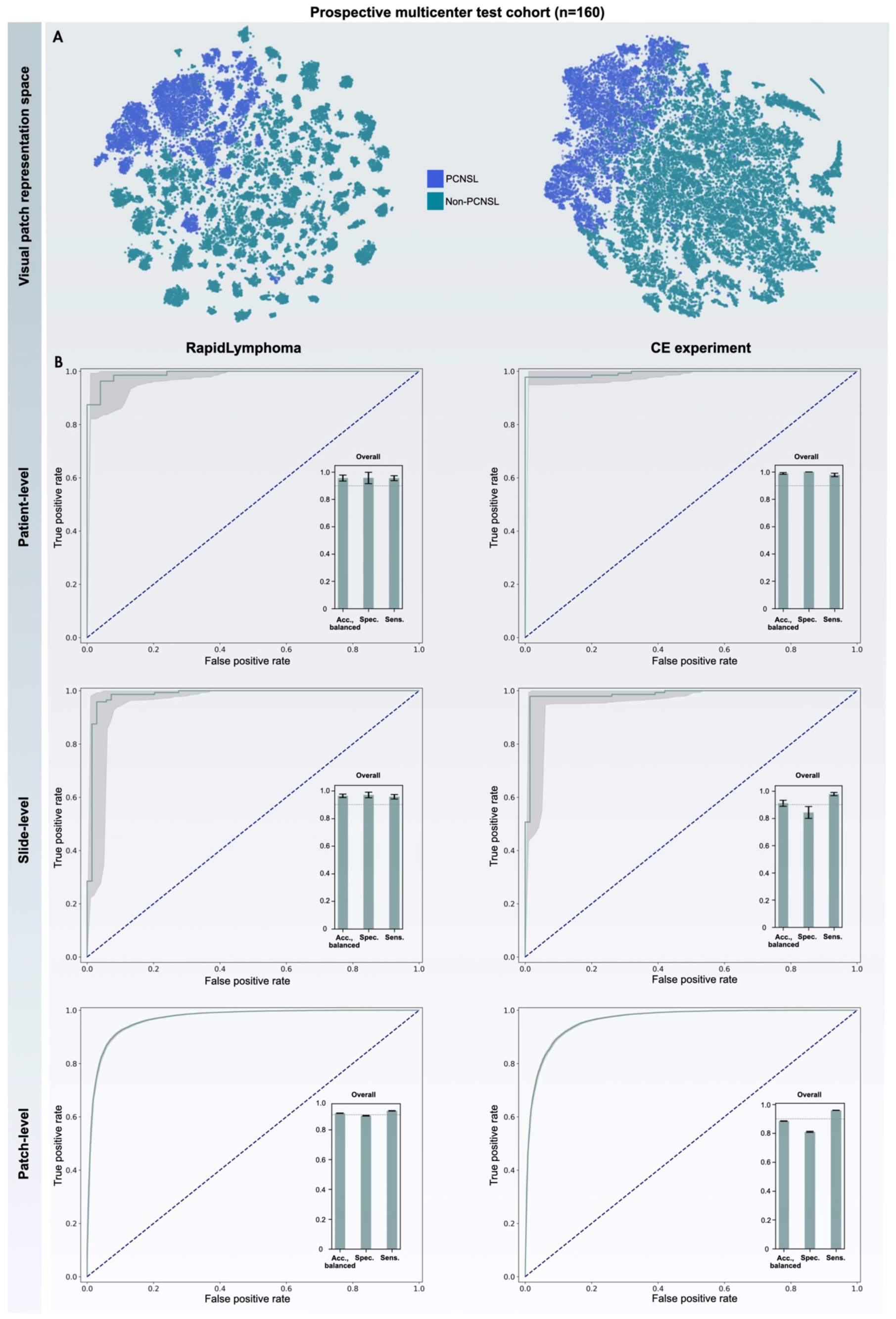
Ablation Study of RapidLymphoma and Cross-entropy Experiment in Prospective Multicenter Test Cohort. This Figure evaluates the performance of the RapidLymphoma model across patient-level, slide-level, and patch-level predictions within the prospective multicenter test cohort. It compares self-supervised learning with cross-entropy (CE) supervised learning as part of an ablation study. **A.** presents t-SNE (t-distributed stochastic neighbor embedding) visualizations of the model’s embedding space for PCNSL (blue) and non-PCNSL (cyan) cases. These visualizations clearly distinguish between the two classes, with PCNSL cases clustering separately from non-PCNSL cases. The RapidLymphoma’s self-supervised approach on the left exhibits more distinct and tighter clustering of PCNSL and non-PCNSL cases compared to the CE-supervised model, suggesting that the self-supervised method is more effective in capturing the underlying structure of the data and achieving better sub-clustering. **B.** contains receiver operating characteristic (ROC) curves and associated performance metrics across patient-, slide-, and patch-level analysis. The rows evaluate predictions on different hierarchical levels. While the CE-supervised method performs well, RapidLymphoma generally achieves better clustering and sub-clustering, as evidenced by the t-SNE visualizations and better results on slide-level predictions used for intraoperative decisions.

RapidLymphoma achieved a balanced accuracy of 97.81% ± 0.91 (95% CI: 95.96-99.29) at the patient- level with an AUROC at 99.70% ± 0.27 (95% CI: 99.01-100.00) (Figure 7B). It identified 25 true positive PCNSL cases and 129 true negative non-PCNSL cases, with 6 false negatives and no false positives. The CE performed slightly better on patient-level, with a balanced accuracy of 98.90% ± 6 (95% CI: 97.45-100), with an AUROC of 99.41% ± 0.40 (95% CI: 98.46-100.00), lower than RapidLymphoma’s performance. CE experiment showed 25 true positive PCNSL cases and 132 true negative non-PCNSL cases, with 3 false negatives and no false positives. Slide-level analysis revealed similar trends. RapidLymphoma achieved a balanced accuracy of 97.21% ± 1.11 (95% CI: 94.71-99.03) and an AUROC of 98.66% ± 1.08 (95% CI: 96.11-99.96), correctly identifying 138 PCNSL slides and 68 non-PCNSL slides, with 6 false negatives and 1 false positive (Figure 8B). The CE experiment showed lower performance with a balanced accuracy of 91.04% ± 2.35 (95% CI: 86.43-95.37) and AUROC with 98.57% ± 0.78 (95% CI: 96.71-99.82), identifying 141 PCNSL slides and 58 non-PCNSL slides, with 3 false negatives and 11 false positives (Figure 8B).

## Discussion

PCNSL is a rare tumor entity of the central nervous system with an incidence rate under 5%.^1,17,18^ Challenges arise from similar clinical, imaging, and pathological features, such as adult-type diffuse gliomas, metastasis, or meningiomas, yet their treatment approaches differ significantly.^1,19–24^ Therefore, tissue sample analysis obtained by biopsy is the gold standard for diagnosing PCNSL, but it poses several challenges, especially in intraoperative settings with limited sample size.^2,6^ Conventional diagnostic methods, such as frozen section and cytology smear analysis, are time- and tissue-consuming and have shown a wide range of accuracy rates, varying between 30.8% and 89.6%.^4,5,7,25,26^ The critical importance of distinguishing PCNSL from other CNS entities intraoperatively, coupled with the limitations of current intraoperative diagnostic methods, underscores the need for consistent and streamlined intraoperative diagnostic tools, especially given the significance of immediate surgical decision-making and subsequent treatment planning. SRH, as the foundational imaging technique, offers several advantages over conventional histological methods, including rapid imaging of fresh, unprocessed tissue samples within minutes and the ability to provide detailed histomorphological information without staining or tissue processing.

The development of RapidLymphoma, our AI-based SRH image analysis tool, addresses the challenges by enabling rapid and accurate intraoperative detection of PCNSL and differentiation from other neoplastic/no-neoplastic CNS entities. Our results demonstrate strong and reliable performance, non-inferior to conventional methods such as frozen section, compared to the final histopathological diagnosis, and are valid to assist surgeons and pathologists intraoperatively. Notably, our system can provide a diagnosis with visual feedback within three minutes, significantly improving over the 20-30 minutes typically required for frozen section analysis (Figure 5).^25,26^ The overall balanced accuracy of over 97% at the patient- and slide-level in our prospective clinical trial demonstrates the robustness of RapidLymphoma in real-world clinical settings. Furthermore, RapidLymphoma demonstrated balanced accuracy rates of over 94% in differentiating typical differential diagnoses such as diffuse gliomas with IDH-wildtype status and various metastases, as demonstrated in the additional test cohorts (Figure 6). This high level of accuracy rates, maintained across various independent test cohorts, different institutions, surgical procedures, and patient demographics, suggests that our pipeline can be reliably integrated into the intraoperative neurosurgical SRH workflow (Supplemental Figure S4 and S5). It underscores the potential of RapidLymphoma to impact clinical decision-making, especially in cases critical for immediate intraoperative surgical strategy and subsequent treatment planning. Our ablation study compared RapidLymphoma with a cross-entropy-based supervised learning approach, providing valuable insights into the advantages of our self-supervised learning strategy without labeling. RapidLymphoma was superior to the CE experiment in cases of visual patch representation embedding with improved clustering of similar patches and demonstrated higher accuracy on slide- and patch-level (Figure 6 + 8 and Supplemental Figure S6). The capabilities of clustering similar patches closer together in the visual representation space of the self-supervised approach enable improved interpretability and meaningful semantic understanding of shared, comparable, and non-similar histomorphological image features. Histologically, PCNSL relies on different discernable features such as noncohesive, monomorphous cell patterns with high nuclear-to-cytoplasmic ratio and angiocentric growth patterns.^6,7^ In general, glial and metastatic CNS tumors remain the critical and challenging differential diagnosis scenarios in conventional histopathologic H&E diagnostics due to similar and overlapping histomorphological features as PCNSL, particularly in preoperative corticosteroid-treated patients with increased cell apoptosis and tissue samples with infiltrative patterns with reactive astrocytes, leading to misdiagnoses and, therefore, further processing for immunohistochemistry and other molecular methods to detect typical PCNSL-associated markers.^6,7^

These overlaps are also challenging for human-based SRH image analysis, where previous work has shown that, there are major difficulties when it comes to distinguishing PCNSL, particularly from metastatic carcinoma.^27^

In our prospective cohort, we identified six false positive and only one false negative case at the slide-level (Figure 4D). Visual heatmaps revealed RapidLymphoma’s predictions with histomorphological similarities between most patches within these slides (Supplemental Figure S2 and S3). The false positive cases comprised two metastatic adenocarcinomas from the lung and four glioblastomas, IDH-wildtype CNS WHO grade 4. These misclassifications can be attributed to similarities in high nuclear-to-cytoplasmic ratios, high chromatin content, and monomorphic cell patterns, resulting in higher pixel intensities resembling those of PCNSL, visible in these SRH images. Also, these misclassifications were correlated with lower SRH image quality, particularly detectable in small stereotactic samples. Even when most diagnostic patches are correctly pre-classified as tumor, since it is a more straightforward classification task, the tissue sample size may be so small that noise from squeezing artifacts or blood contamination can hinder confident patch-based non-PCNSL classification. Although the number of diagnostic patches and the associated tissue sample size also play a role in AI-based SRH image analysis, significantly less tissue is generally required for accurate diagnosis than in conventional intraoperative methods, which is critical in stereotactic-guided biopsies.^16^ The histomorphological similarities described above could also be found in the two additional test cohorts, focusing on classifying only glial and metastatic CNS tumors. One case in the prospective clinical trial resulted in a false negative predicted case that revealed an infiltrative pattern with reactive astrocytes, typically represented in glial tumors, and demonstrated only in a non-representative number of true-positive patches, PCNSL-specific patterns. This lower rate of false negatives classified SRH whole-slide images further underscores the model’s high specificity, which is crucial for accurate intraoperative decision-making, especially in cases with limited tissue samples. High specificity is considerably more critical than sensitivity when making correct intraoperative surgical decisions, especially during stereotactic-guided biopsies, as the treatment strategies for PCNSL and non-PCNSL entities differ significantly. The importance of specificity is further enhanced when combined with a high slide-level confident probability for PCNSL and visual feedback provided by RapidLymphoma, allowing surgeons to make more informed decisions.

Nevertheless, RapidLymphoma enables visual disclosure of PCNSL-typical histomorphological features, including high chromatin content, high nuclear-to-cytoplasmic ratio, mono- to pleomorphic cell structures, angiocentric growth patterns, and increased reticulin fiber structures. The misclassifications underscore the challenges in differentiating PCNSL from other malignancies with similar cellular characteristics, especially in cases with limited sample size or quality. However, RapidLymphoma’s ability to highlight key PCNSL features demonstrates its potential as a valuable tool in intraoperative diagnosis when used with other clinical and pathological information.

In our pipeline, continuous expansion of the training dataset to include a broader range of CNS pathologies and unusual presentations of PCNSL is simple to accomplish and could further enhance diagnostic capabilities.

Moreover, our developed AI-based pipeline represents a simple self-supervised learning implementation not limited to the neurosurgical field; therefore, it is eligible for other AI-based SRH image classification tasks.

### Limitations

Several conditions limit this study. While the prospective international multicenter design is a strength, the relatively small number of PCNSL cases in this cohort reflects this condition’s rarity, potentially biasing the specificity of the prospective cohort. Another limitation to consider is the inherent "black box" nature of deep learning models like RapidLymphoma, which makes it challenging to understand the exact decision-making process of the model entirely. However, the use of visual heatmaps as an interpretability tool, while not providing a complete explanation of the model’s decision-making process, does offer clinicians valuable insights into which regions of the SRH images are most influential in RapidLymphoma’s predictions during surgery. Furthermore, it would be valuable to examine further how this treatment influences AI-based SRH image classification, particularly in PCNSL. However, our training and test set included tissue samples affected by preoperative corticosteroid therapy and performed well, reflecting real-world scenarios. Another limitation is that our study developed an AI-based image classification task for SRH limited to institutions with an SRH imager. However, our developed AI-based pipeline for RapidLymphoma is not limited to SRH images. It is also eligible and easily changeable for any other input data derived from, e.g., hematoxylin and eosin, immunohistochemistry, or other histopathological images.

In conclusion, RapidLymphoma is a valuable and reliable computer vision tool for intraoperative diagnosis of PCNSL. By leveraging the strengths of SRH and advanced deep learning techniques, this pipeline addresses assistance in current diagnostic approaches, offering rapid, accurate, and interpretable intraoperative diagnoses. The ability to provide a diagnosis with visual feedback within just three minutes while maintaining accuracy non-inferior to conventional frozen section analysis is a feature that can significantly streamline neurosurgical procedures and could lead to improved patient outcomes through more informed and timely surgical decision-making.

## Supporting information

Supplemental Table S1 - Patient-Level - Prospective Cohort

Supplemental Table S2 - Slide-Level - Prospective Cohort

Supplemental Table S3 - Patch-Level - Prospective Cohort

Supplemental Table S4 - Training Cohort

## Data Availability

All data produced in the present study are available upon reasonable request from the authors. The code used in this work will be made available after it is accepted for publication in a peer-reviewed journal.

## Authorship

D.A.O. and D.R. conceptualized and designed the study. D.R. developed the AI models, performed the computational analyses, and made the visualizations. D.R. wrote the original manuscript draft. D.R., N.M., A.S., J.M., N.K.G., T.C.H., A.C., C.J., X.H., V.N., A-K.M., G.F., M.I.R, D.Ru. L.K., G.W., D.A., J.G.G., and D.A.O. collected tissue for SRH imaging and processed the SRH images. M.S., T.R-P., T.S., and A.A-S. processed and analyzed tissue for frozen section and final histopathological diagnosis. All authors wrote the manuscript with input from all authors. All authors contributed to the manuscript review and editing and approved the final version.

## Data availability

Due to identifiable patient data, raw SRH image data cannot be shared publicly. However, de-identified SRH patch images and their corresponding labels can be requested from the corresponding author, subject to appropriate data use agreements. Our AI-based pipeline code repository for RapidLymphoma is publicly available after acceptance and publication.

## Acknowledgments

We thank the neurosurgical teams at New York University, the University of Cologne, Michigan Medicine, and the Medical University of Vienna for supporting patient recruitment and tissue collection. The computational requirements for this work were supported in part by the NYU Langone High Performance Computing Core’s resources and personnel.

---

**Supplemental Figure S1.**
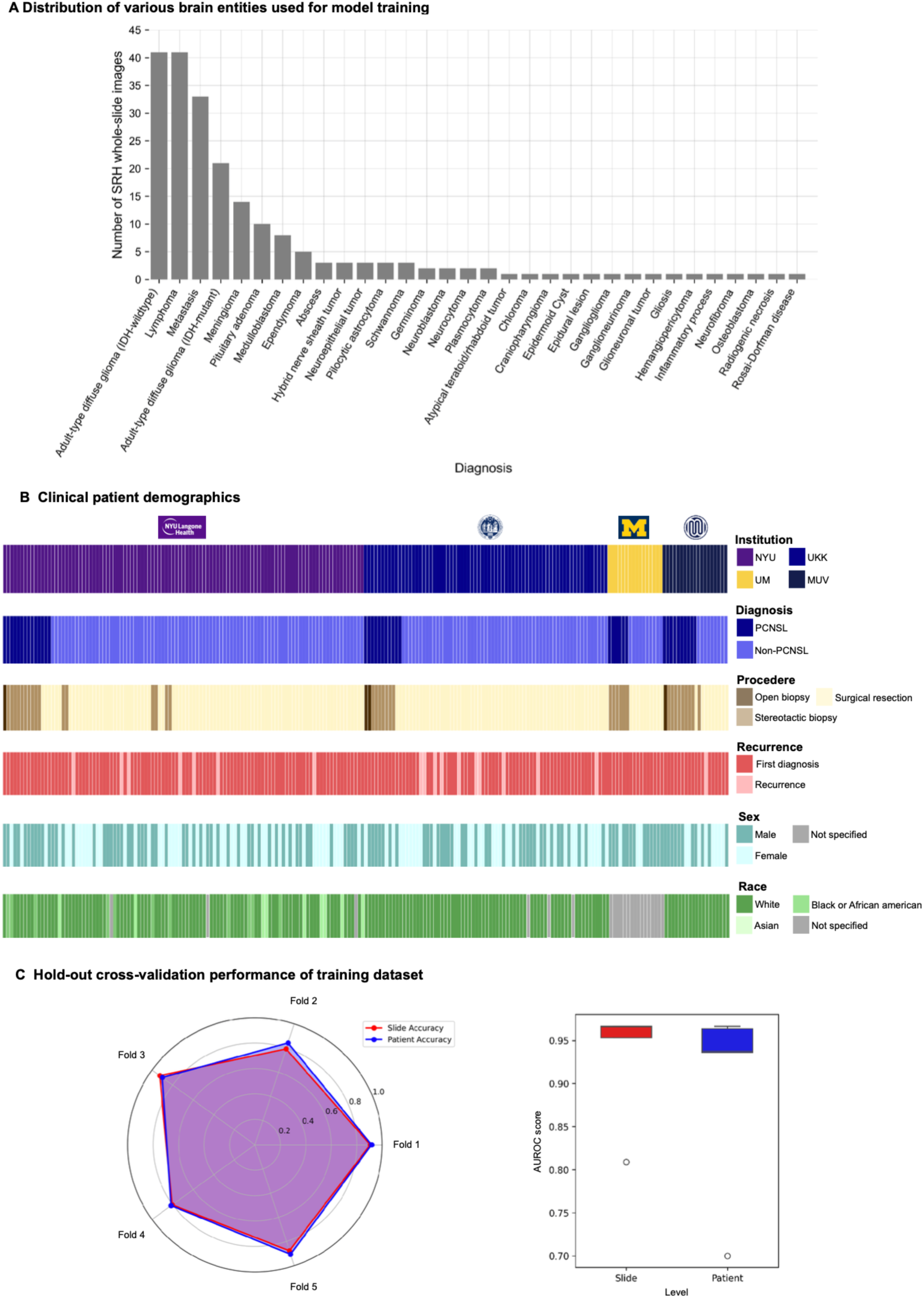
Dataset Composition and Model Performance Evaluation. An overview of the dataset composition and model performance used for training in this study is presented. **A.** illustrates the distribution of various CNS entities included in the model training, with the highest number of cases attributed to the typical differential diagnosis of primary CNS lymphoma (PCNSL). **B.** provides a detailed breakdown of the clinical patient demographics across different institutions, denoted by distinct colors: New York University (NYU), University of Cologne (UKK), Michigan Medicine (UM), and Medical University Vienna (MUV). Corresponding demographics are represented for diagnosis, procedure, recurrence, sex, and race. **C.** evaluates the hold-out cross-validation performance of the training dataset. The chart on the left compares slide-level and patient-level accuracy across five folds. With the self-supervised learning strategy, slide and patient accuracy perform well in all folds. On the right is a bar chart showing the AUROC scores for slide-level and patient-level predictions, indicating high predictive performance, with scores approaching 0.95 for both levels, underscoring the robust performance and diverse representation within the dataset used for model training.

**Supplemental Figure S2.**
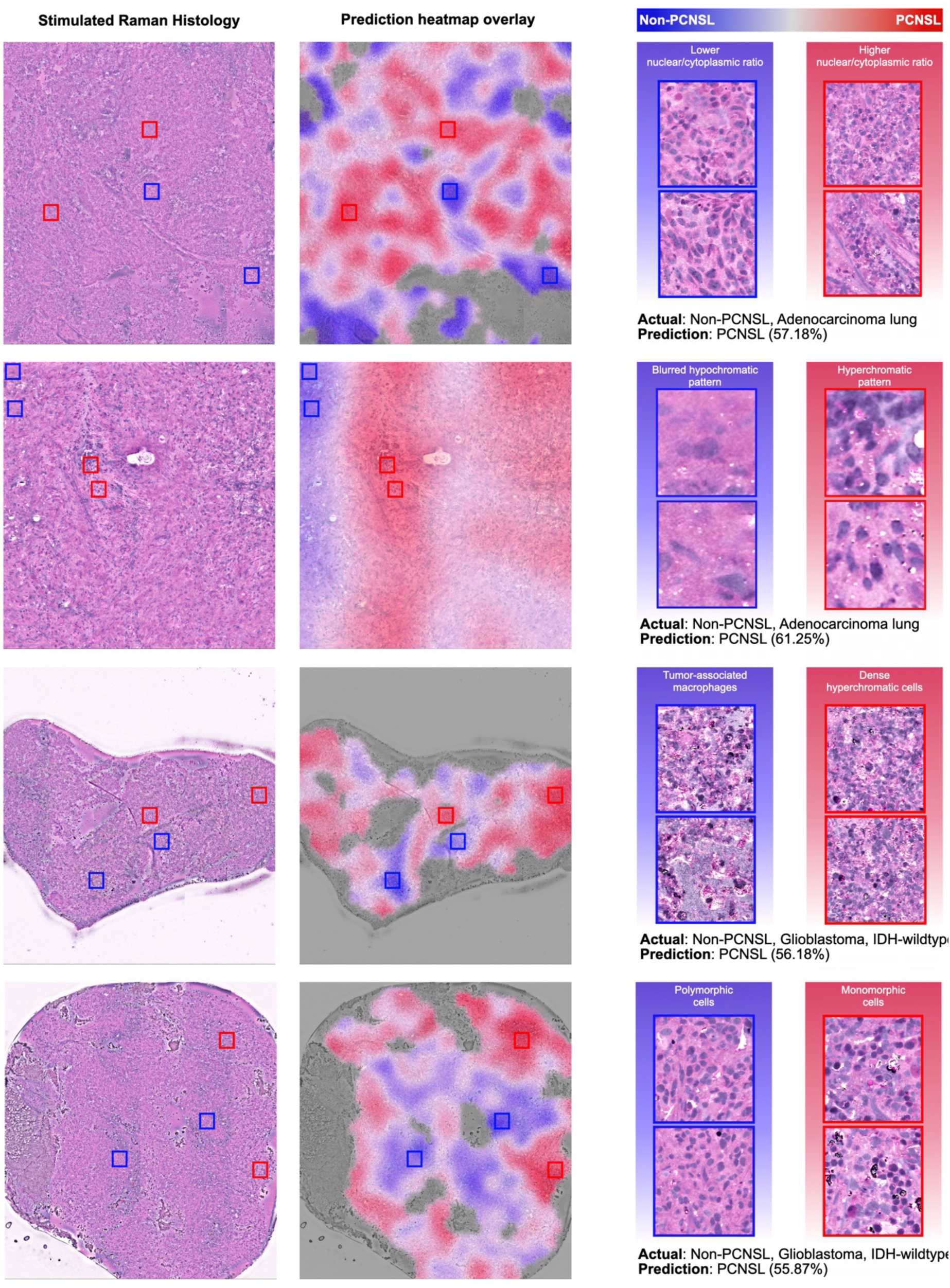
Misclassified False Positive Whole Slide Images from the Prospective Testing Cohort. This figure displays four false positive whole slide images from the prospective test cohort, illustrating the challenges and complexities in distinguishing between PCNSL and non-PCNSL tissues. On the left the Stimulated Raman Histology (SRH) whole slide images, RapidLymphoma’s prediction heatmap overlays in the middle, and detailed patch comparisons from the marked red and blue squares indicating the model’s predicted regions as PCNSL and non-PCNSL, respectively on the right. For each misclassified case, the actual diagnosis and the model’s incorrect prediction are noted alongside the softmax scores on slide-level. The heatmaps use color gradients to represent the model’s confidence levels, with red indicating high confidence for PCNSL, blue for non-PCNSL, and white/grey for low confidence. A detailed analysis of the patches from the misclassified slides shows similar features across the two classes, e.g., high nuclear/cytoplasmic ratio or monomorphic cell patterns. Noisy images, such as from small biopsy tissue samples (Image 3 and 4), also influence prediction behavior.

**Supplemental Figure S3.**
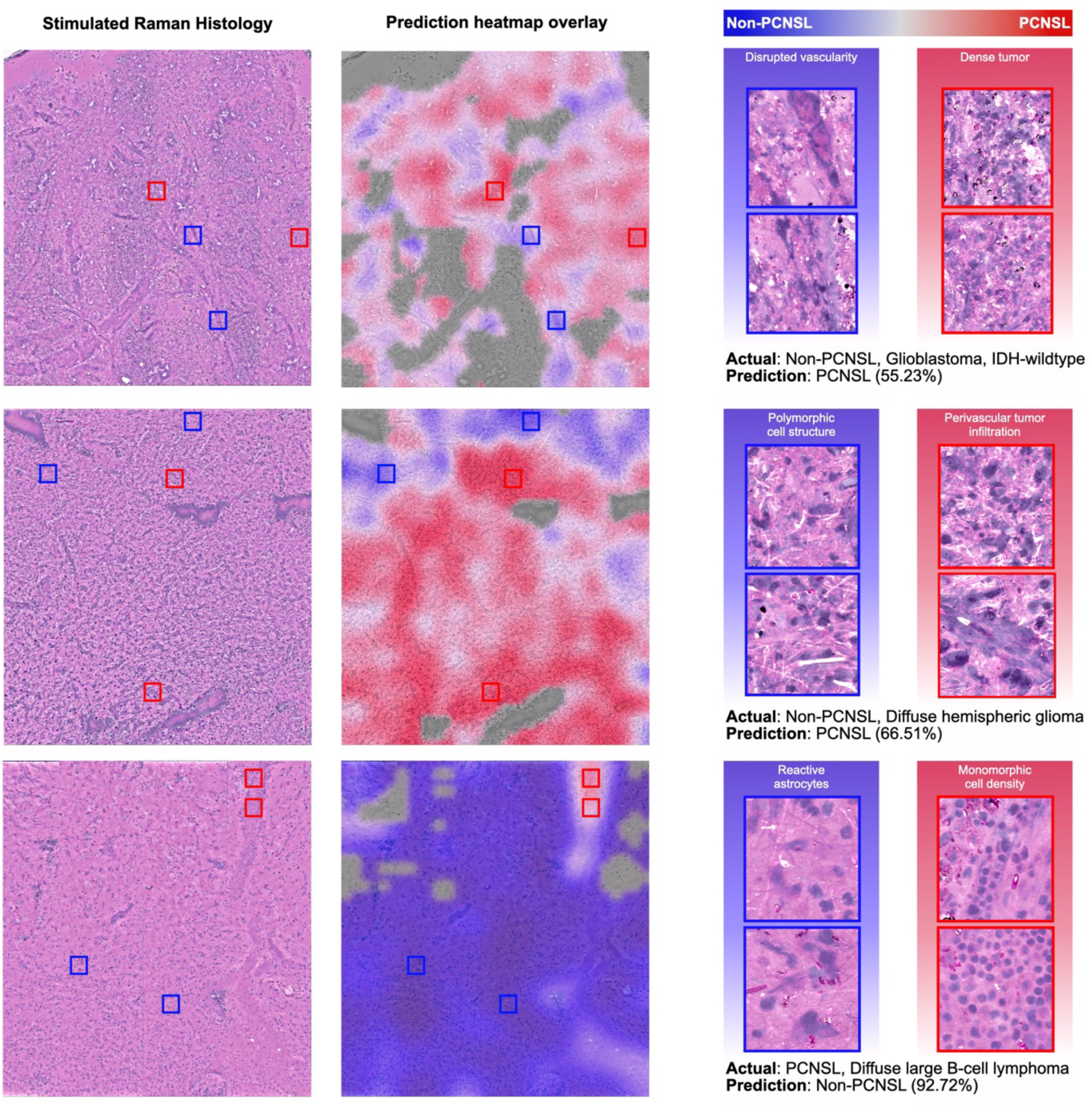
Misclassified False Positive and Negative Whole Slide Images from the Prospective Testing Cohort. The figure presents three false positives and one false negative predicted SRH whole-slide images alongside RapidLymphoma’s prediction heatmaps and detailed patch comparisons. For each misclassified case, we provide the actual diagnosis, the model’s prediction, and slide-level softmax scores. Heatmaps use color gradients to visualize the model’s confidence levels, with red and blue indicating high confidence for PCNSL and non-PCNSL, respectively. Analysis of the detailed patches reveals similar features across classes, such as high nuclear/cytoplasmic ratios or monomorphic cell patterns, as well as image quality contributing to classification difficulties. Notably, the false negative SRH whole-slide image demonstrates infiltrative character with a high density of reactive astrocytes typically seen in glial tumors.

**Supplemental Figure S4.**
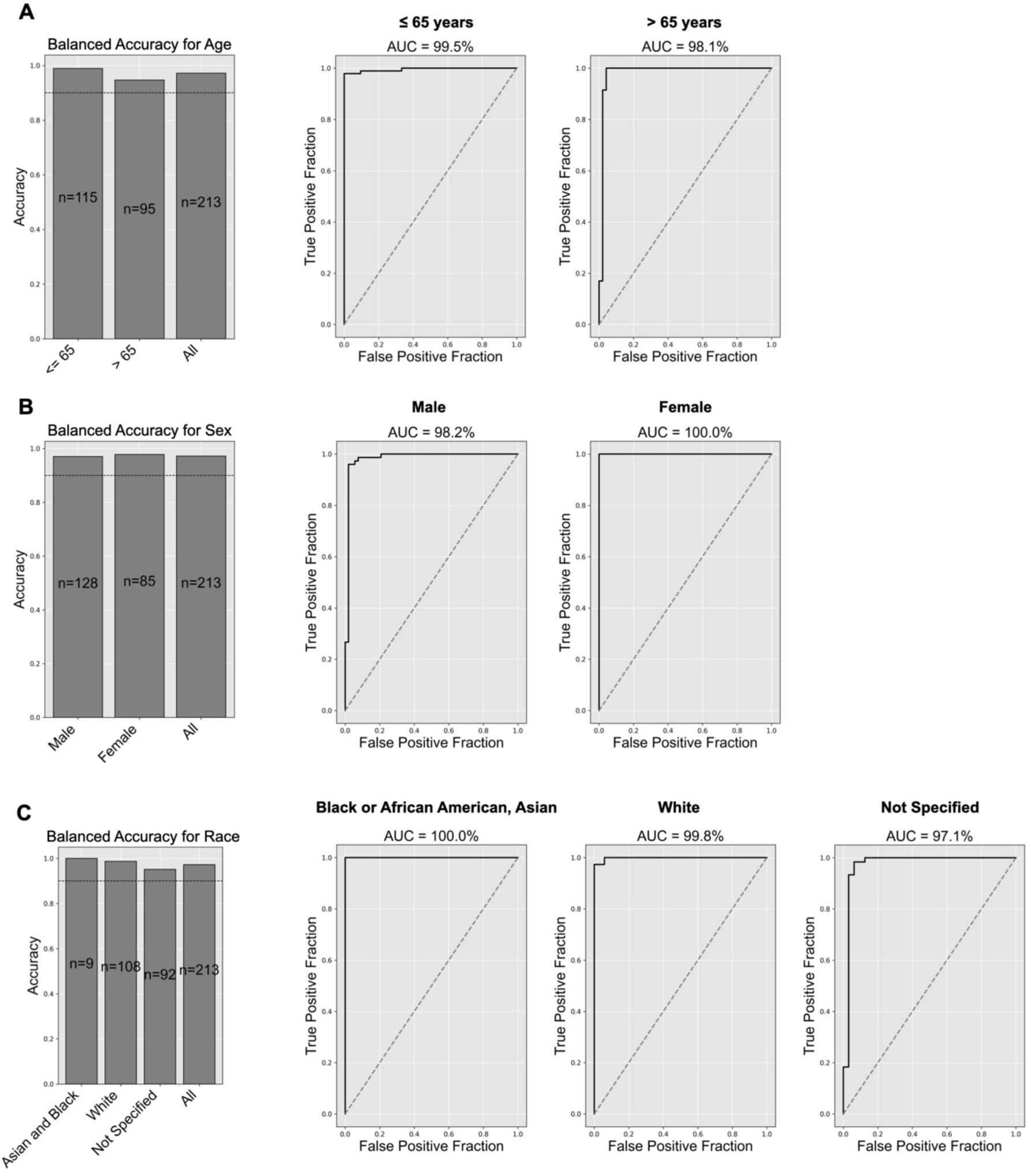
Patient Demographic Sub-analyses of the Prospective Multicenter Test Cohort on Slide-Level. RapidLymphoma model’s performance across different demographic subgroups based on age, gender, and race is shown, presenting balanced accuracy and ROC curves for each category. **A.** focuses on age groups, showing highly balanced accuracy and ROC curves with high confidence and specificity for individuals aged ≤ 65, > 65, compared to the entire cohort, indicating robust classification across different age groups. **B.** examines gender-based performance, demonstrating highly balanced accuracy and ROC curves with strong sensitivity and specificity for both male and female subgroups. **C.** analyzes performance by race, with highly balanced accuracy and ROC curves for Asian and Black or African American, White, and those not specified, reflecting effective classification and high confidence across racial subgroups. Overall, it confirms the high and balanced accuracy of the RapidLymphoma model across various demographic categories, underscoring its reliability and generalizability in clinical applications.

**Supplemental Figure S5.**
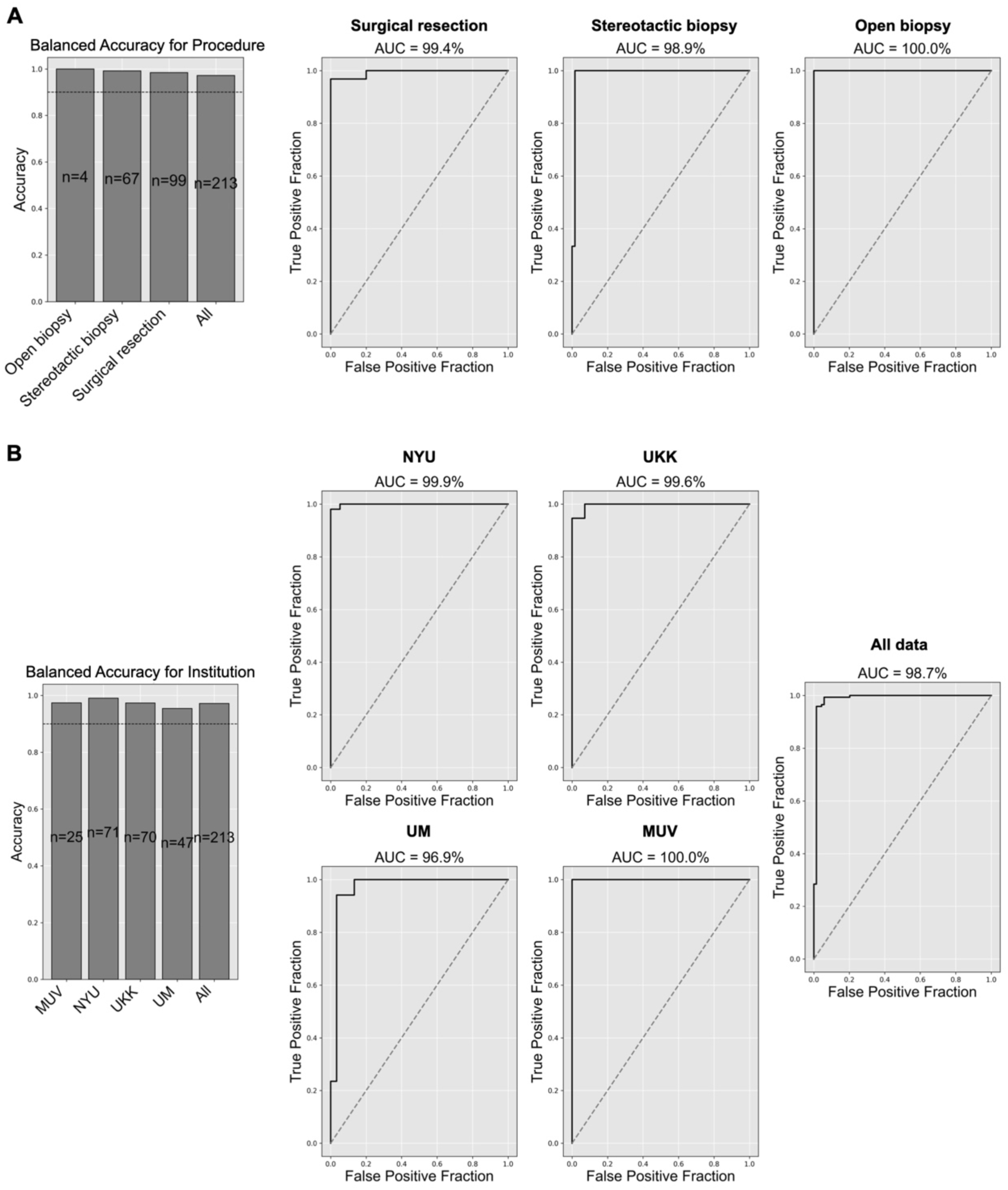
Patient Demographic Sub-analyses of the Prospective Multicenter Test Cohort on Slide-Level. **A.** focus on different medical procedures, including open biopsy, stereotactic biopsy, and surgical resection. The balanced accuracy and ROC curves for each procedure type show high confidence, specificity, and sensitivity, indicating robust classification performance across various surgical methods. **B.** evaluates the model’s performance across different institutions: New York University (NYU), University of Cologne (UKK), Michigan Medicine (UM), and Medical University Vienna (MUV) and for all data combined. The balanced accuracy and ROC curves for each institution demonstrate high confidence, specificity, and sensitivity, reflecting the model’s consistent accuracy and reliability regardless of the institution, underscoring its robustness and generalizability in diverse clinical settings.

**Supplemental Figure S6.**
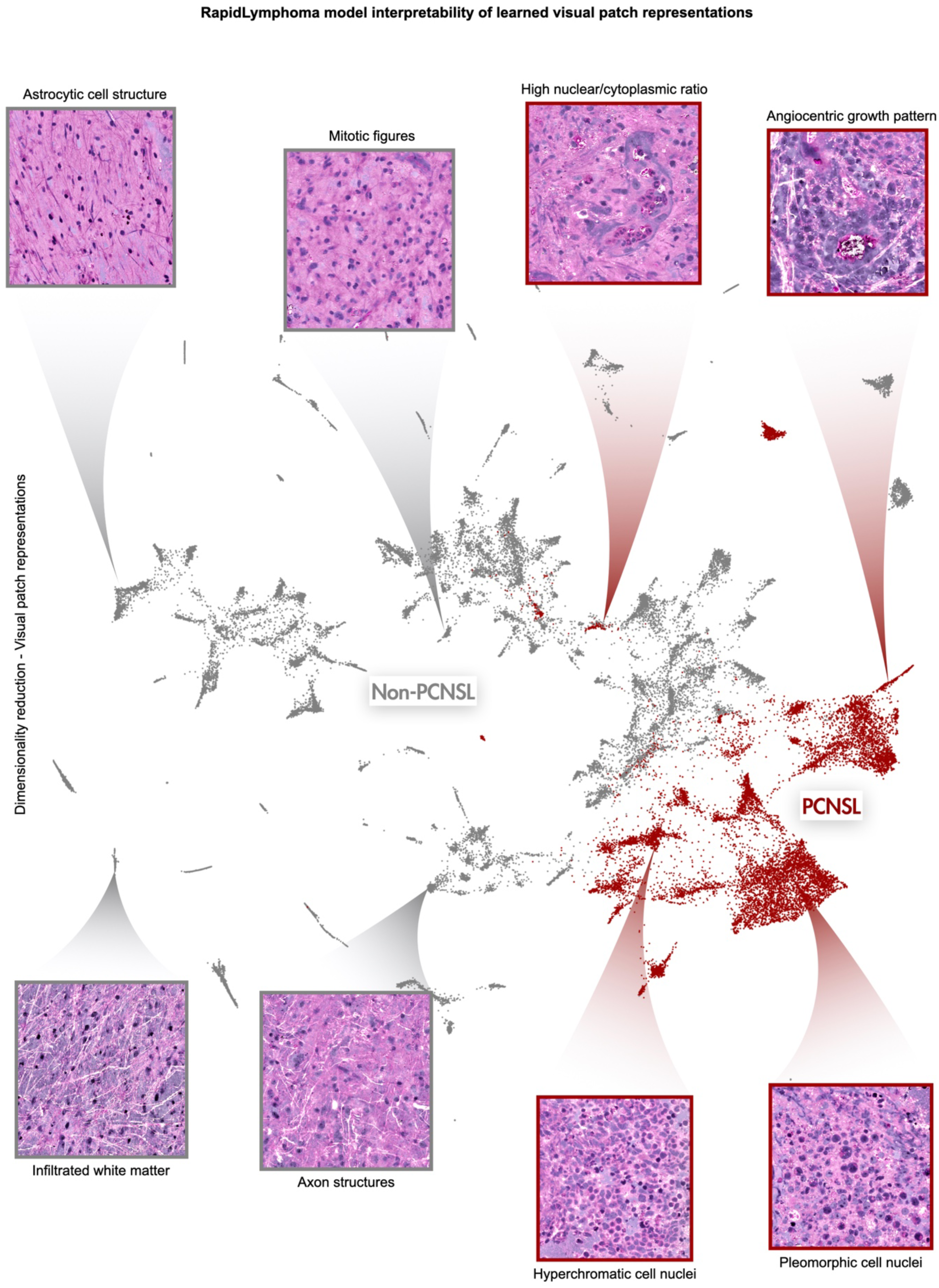
Interpretability of RapidLymphoma Model’s Learned Visual Patch Representations. This figure illustrates the interpretability of the RapidLymphoma model by highlighting the visual patch representations that the model has learned to distinguish between primary CNS lymphoma (PCNSL) and non-PCNSL tissues. The central plot represents the embedding space of the visual patches of the prospective multicenter test cohort, visualized using PaCMAP (Pairwise Controlled Manifold Approximation Projection). Each point in the plot corresponds to a patch from the SRH images, with points color-coded to indicate whether they belong to PCNSL (red) or non-PCNSL (gray) regions. The clear separation between the two clusters demonstrates the model’s ability to differentiate between these two classes based on learned features. Surrounding the plot are representative patches that exemplify key histological features the model identifies, following a representational and histomorphological direction from left to right.

